# Facilitators and barriers in the use of the electronic consultation register for Integrated Management of Childhood Illness in the health district of Toma, Burkina Faso: Perspectives of health care providers

**DOI:** 10.1101/2024.03.29.24305086

**Authors:** Ngangue Patrice, Saouadogo Issaka, Kaboré Soutongnoma Safiata, Mbang Massom Douglas, Bationo Nestor, Josiane Seu, Birama Apho Ly

## Abstract

**Background:** In collaboration with the Ministry of Health and Public Hygiene (MHPH) of Burkina Faso (BF), the Foundation Terre des Hommes (Tdh) has developed the Integrated e-Diagnostic Approach (IeDA) project in BF since 2010 to strengthen the health system by digitalizing medical protocols, improving the quality of services and using data. We sought to identify and analyze the barriers and facilitators of using the electronic clinic registry (ECR) for the integrated management of childhood illness (IMCI) by healthcare providers (HCPs) in the health district of Toma, BF.

**Methods:** We conducted a descriptive and exploratory qualitative study. In-depth individual interviews were conducted with thirty-five (35) HCPs in the health district of Toma, BF, from the 1^st^ to the 30^th^ of December 2021. Thematic analysis of qualitative data according to the Braun & Clarke model was performed using NVivo 12 software and arranged along a social-ecological model.

**Results:** Our findings revealed that HCPs play an essential role in using ECR for IMCI. Many key facilitating factors have emerged regarding the use of IMCIs in primary health care (PHC) facilities, such as positive perceptions of the ECR, firm commitment and the involvement of HCPs, stakeholder support, collaborative networks with implementing partners, convenience, privacy, confidentiality and client trust, experience and confidence in using the system, and the satisfaction, motivation and competency of staff. In addition, the easy diagnosis offered by the ECR and the training of HCPs increased the acceptance and use of the ECR. Regarding barriers, HCPs complained about the tablet’s slowness, recurrent breakdowns, and increased workload.

**Conclusion:** This study revealed that ECR has excellent potential to improve the quality of care and, in turn, reduce maternal and infant mortality. Although the satisfaction of the HCPs with the tool is positive, the actors of the Foundation Tdh, in collaboration with the MHPH, must work to optimize the application’s performance and reduce breakdowns and delays during consultations. This will allow the deployment of ECR in all BF health districts.

## Introduction

Information and communication technologies (ICTs) have been essential in various sectors for decades, providing numerous professional advantages (Al-Ansi, Garad, & Al-Ansi, 2021; Lidström & Hemmingsson, 2014). The interest in ICTs has recently intensified in the health sector (Alot Federico, 2017). Its introduction and implementation in the developed world have shown its effectiveness in providing quality care at acceptable and tolerable costs for the population (Bali, 2019).

In developing countries, adopting ICTs in health systems is necessary to improve people’s health and well-being (Shao, Fan, Huang, & Chen, 2022). To reduce maternal and child mortality in Burkina Faso (BF), the Integrated Management of Childhood Illness (IMCI) strategy, an algorithm that systematically guides health professionals in the clinical evaluation process of children under five years who come for a consultation, was adopted in 2003 (Gera, Shah, Garner, Richardson, & Sachdev, 2016; Traoré et al., 2019). As healthcare is one of its priority areas of intervention, the Foundation Terre Des Hommes (Tdh) has supported the Ministry of Health and Public Hygiene (MHPH) of BF in improving the health of children under five years of age (5) through technological innovation. To improve compliance with the guidelines of the IMCI protocol in health centers, the Foundation Tdh has developed the *Integrated e-Diagnostic Approach* (IeDA) strategy since 2010 (LSHTM, 2018). This strategy aims to improve the implementation and use of the IMCI. IeDA is a telehealth solution for reducing child mortality through digitalizing medical protocols, improving quality, enhancing data use, and continuously training agents. It also relies on the Electronic Consultation Record (ECR) (Blanchet, Sanon, Sarrassat, & Somé, 2022). This software guides health professionals throughout the consultation process based on the IMCI by recording achievements and rating the adherence of healthcare providers (HCPs). For this instance, the Bill and Melinda Gates Foundation partnered with the Foundation Tdh to implement a project to strengthen the health system regarding maternal health, malaria, and universal health coverage. This project was implemented in the health district of Toma in BF in collaboration with the MHPH. The applications used in the framework of the maternity ECR were developed with the help of several departments within the MHPH National Malaria Control Program, the Directorate of Statistics Sector (DSS), the Directorate of Health and Family, the Directorate of Health Information Systems, the Regional Directorate of Health and the district of Toma, learned medical societies in BF and the Foundation Tdh. It was then deployed in the health district of Toma in 2020 to be used by healthcare providers (HCPs) in the Health and Social Promotion Centers (HSPCs). After a few years of implementing the IeDA, a realistic evaluation was conducted to describe the project effects and users’ perceptions and analyze factors that influenced the implementation (Blanchet et al., 2022). However, no study has explored the barriers and facilitators that HCPs encounter in adopting and using this new tool. This study aimed to assess these barriers and facilitators from the HCPs’ perspectives in using the ECR for IMCIs in the health district of Toma, BF.

## Methods

This study used a qualitative descriptive and exploratory design. Verbal data were collected to ensure an interpretative approach. The study was conducted in the health district of Toma from 1^st^ to 30^th^ December 2021. The reporting of this study was guided by the Consoli dated Criteria for Reporting Qualitative Research (COREQ) guidelines (Tong, Sainsbury & Craig, 2007).

### Participants and recruitment

The study participants were HCPs from Toma’s health district who used the ECR for IMCIs. The recruitment of participants was based on the following inclusion criteria: 1) were HCPs in the health district of Toma; 2) were involved in consultation and patient management activities; 3) had received training on the use of the ECR for IMCI; 4) were present on the day of the visit of the investigators; and 5) provided free and informed consent to participate in the study without coercion. In addition, any HCP who expressed a desire to refrain from participating after completing the questionnaire was excluded from the study.

### Data collection

We conducted individual semi structured interviews to collect primary data from the 1^st^ to the 30^th^ of December 2021. HCPs working in the HSPC in Toma’s health district were considered for interviews. For this purpose, an interview guide was developed and used to collect participant data. The themes developed in the interview guide focused on questions related to the objectives and concepts covered by the research. The data collection tools were first tested in the health district of Tougan and validated by consensus between the data collectors and the principal investigator. The latter district has the same characteristics as the Toma district. These districts all belong to the Boucle du Mouhoun health region. All interviews were recorded using an audio recording device, and the average length of each interview was forty-five (45) minutes. Additional notes were collected during the interviews using notepads, and a debrief session was held between the data collectors and the principal investigator at the end of each interview.

### Data processing and analysis

The collected data were processed and analyzed using the Braun and Clarke approach (2006). In the first step, we exported the interview recordings to a computer for transcription. Then, the verbatims were exported to Microsoft Word for coding. Finally, the transcripts were read several times to determine their meaning and significance. In **step 2**, all the verbatims were coded according to the themes and categories derived from the data. In **step 3**, the codes were grouped into subthemes to highlight the meaning units. A synthesis was made following this analysis based on similarities and differences. **The fourth step** was dedicated to reading the potential themes developed for refinement. At this stage, some themes were grouped, and others were separated. New themes were created according to the needs of the analysis. In step 5, each theme was definitively renamed and retained for the study. The sixth step involved presenting the results of the thematic analysis of the data. The relevant verbatim extracts from the collected data were used to illustrate the interpretations and results in the discussion.

### Ethical considerations

Administrative authorization was obtained from the local health authorities for this study after approval from the Burkina Faso Health Research Ethics Committee (N°2021-12-283). The objectives, voluntary nature, importance of participation in the research and possibility of withdrawal were explained before each interview. The confidentiality of the information collected was ensured by limiting access to the data only by the investigators and to the needs of the survey.

To guarantee anonymity, each respondent was assigned a unique identification code to conceal their identity. This coding was constructed by considering the gender and seniority of the participants. The results are presented so that participants cannot be identified.

## Results

### Sociodemographic characteristics of the respondents

The study sample consisted of thirty-five (35) participants, twenty-three (23) men and twelve (12) women. Participants’ ages ranged from twenty-seven (27) to forty-three (43) years. Nineteen (19) participants were nurses, seven (07) were midwives, six (06) were mobile health and community hygiene workers, and three (03) were birth attendants. The professional experience of these questionnaire respondents ranged from one (01) to five (05) years. In addition, all the HCPs had experience using the ECR, ranging from one (1) to five (5) years. The sociodemographic characteristics of the respondents are shown in Table 1 below.

**Table 1:**
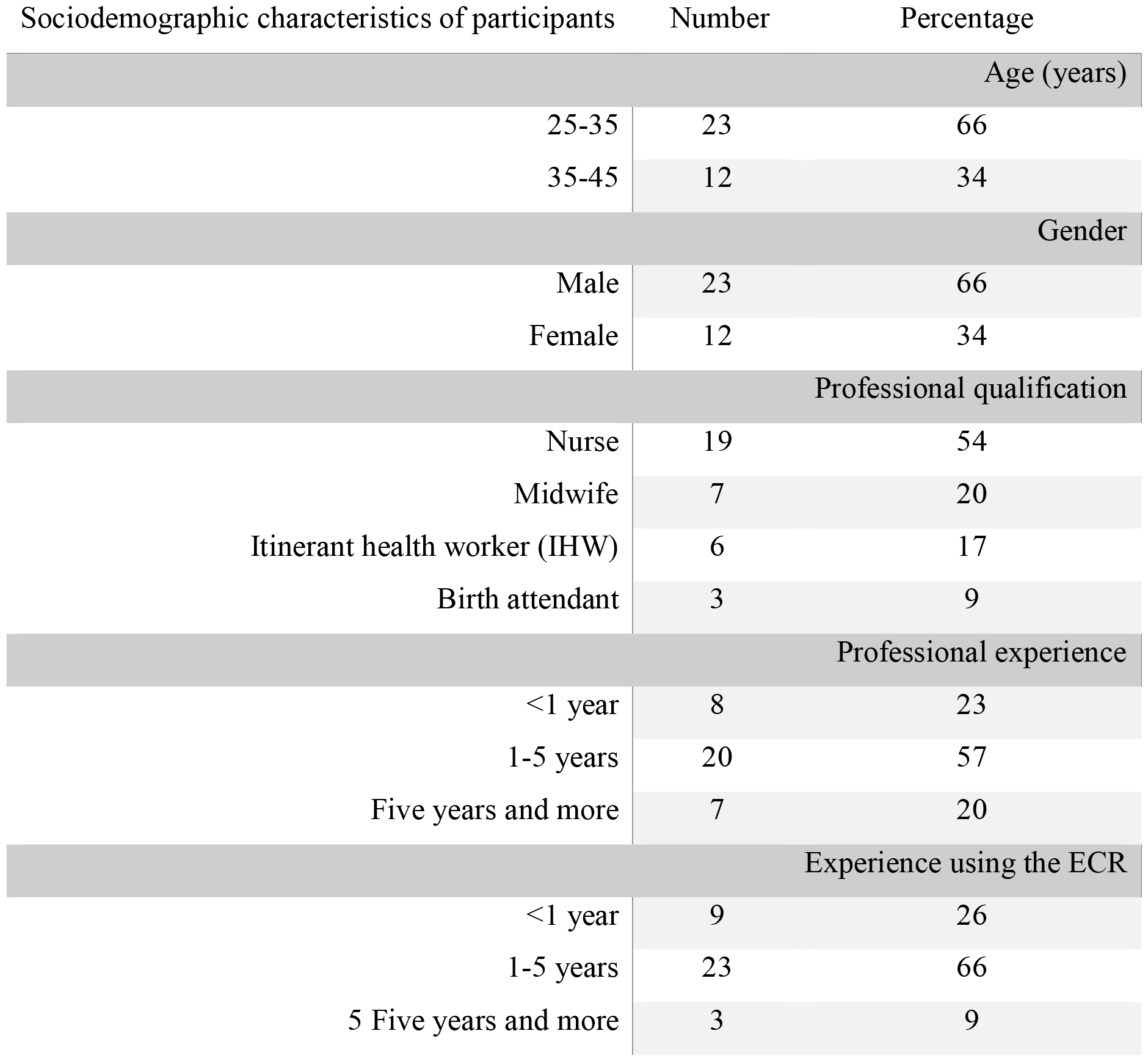
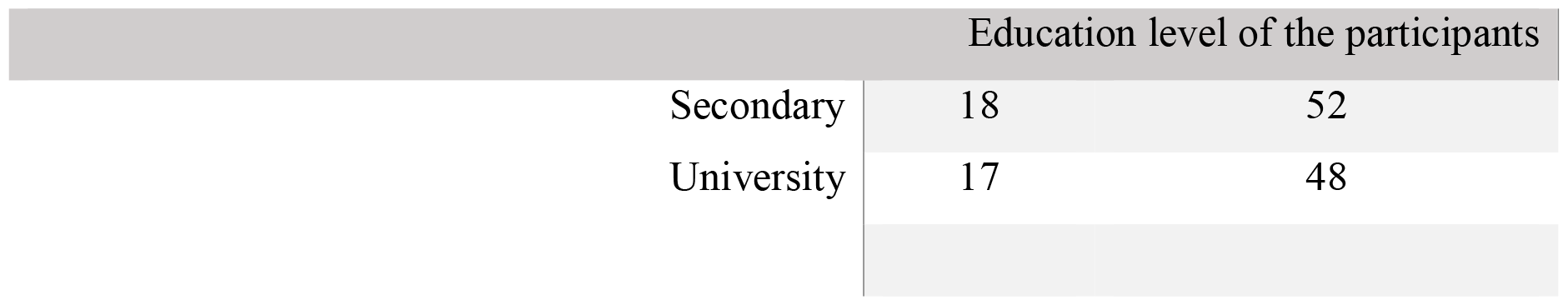
Sociodemographic characteristics of the respondents.

### Factors related to the use of the electronic consultation register

The findings revealed that many factors contributed to ECR use. One of these factors is the overall positive perception of the participants regarding ECR.

In this respect, participants felt that ECR was a valuable aid to diagnosis and proper patient management, as illustrated by this patient’s words: *“It is a guide for the health worker. It allows for proper monitoring of women. It also allows the health workers to train themselves”* **(Participant 4)**.

In addition to the participants’ positive perceptions of using ECR, it is also interesting to mention, along with health workers, that ECR allows health workers to follow the clinical process without missing any clinical signs. In this way, ECR is also a facilitating factor. Clearly, after each symptom assessment, the ECR directly proposes a classification without the health worker resorting to his chart book or other management guides (paper version). Moreover, the ECR offers speech bubbles, images, and videos, allowing health workers to identify or recognize signs quickly. Furthermore, the accuracy of ECR is such that at the end of the assessment, the health worker is given a suggested course of action regarding drugs and the posology to be prescribed to the patients. This tool allows health workers to have access to patients’ previous diagnostic history when they go to the health facility for a new consultation. In this sense, a participant reported that:

*“Rationally, it facilitates the registration of the patient, the diagnosis and the treatment, so it facilitates the management”* **(Participant 35)**.

Training staff to use the ECR (either face-to-face or during coaching) at the beginning was a relevant strategy because it increased the support of health staff for its use. In the beginning, the Foundation Tdh, in collaboration with the health districts, organized capacity-building sessions lasting between four (4) and six (6) days to benefit HCPs. This capacity building has given rise to a feeling of doing better to change practices and apply what they have learned during the sessions. Furthermore, during these training sessions, agents received financial compensation at the end of the sessions. This financial compensation increased adherence and created a sense of accountability for t he district management team and the Foundation Tdh. This has contributed to better implementation and acceptance of the strategy. In support of this, the following narrative illustrates: *“Factors that contributed to the success of the ECR included training health workers on the ECR tool”* **(Participant 33)**.

### Barriers to the use of the electronic consultation register

According to the interviewees, technology is the main barrier to using the ECR. Two (2) significant barriers were identified: technological hazards and the increased workload associated with using the ECR.

The first barrier concerns the technical bugs of the application, the slowness, and recurrent breakdowns of the tablet. The use of the tablet and the issue of synchronization led to an additional workload, according to the respondents. Most of the respondents’ comments highlighted feelings of discouragement and frustration that often led to little or no use of the ECR. Thus, specific difficulties affect the quality and effectiveness of ECR use. Among these difficulties is the performance of tablets deployed in health facilities. The Foundation Tdh, in collaboration with the MHPH, deployed large-capacity and high-end state-of-the-art tablets. However, given technological developments, these tablets became obsolete after three years of deployment and could no longer support the ECR and the updated applications. This situation concerned the extensive health facilities that provided data. This helped to explain some of the slowness problems highlighted by the participants. One participant rightly stated that *“The slowness of the ECR can be noted, especially with the Integrated Management of Childhood Illness (IMCI)”* **(Participant 12)**.

The second barrier is the internet network’s poor quality and IT support. Indeed, the poor quality of the Internet infrastructure at the level of the health care facilities led to synchronization problems and, in turn, to a slowing of the application and, thus, of the consultation. This was because the tablet contained important data from previous consultations. This situation worsened when HCPs wanted to carry out online consultations. A participant who shared this opinion said, *“First, there is the slowness; the device sometimes presents a slowness that does not allow the consultation; beyond that, there is the connection”* **(Participant 24)**.

In addition, the IT staff of the Foundation Tdh’s lack of knowledge of the ComCare platform was also a barrier. As ComCare was developed and managed by DIMAGI, an independent partner organization, this explains the project’s IT staff’s need for more mastery of this platform. The DIMAGI was responsible for hosting the ECR data from the beginning of the intervention. However, in this collaboration, DIMAGI only gave Tdh’s IT staff limited access to the platform. This created a dependency and did not facilitate the prompt processing of requests. A concrete example was reported: *“It is the frequent breakdowns that prevent us from using the ECR”* **(Participant 10)**.

There is also a need for more capacity at the district level to respond urgently and effectively to the various requests submitted by healthcare providers. The following statement illustrates this: *“The main factor limiting the use of the ECR is blocking the ECR”* **(Participant 30)**.

## Discussion

Our study aimed to document the barriers to and facilitators of ECR use by HCPs in the health district of Toma, BF. Our results revealed that the ECR is a relevant tool for diagnosing and managing patients. Furthermore, the training of all HCPs in ECR use was also identified as facilitating ECR use. On the other hand, the factors that hindered the use of the ECR were mainly related to the slowness and recurrent breakdowns of the tablet.

The analysis revealed that facilitators contributed to the successful implementation of the ECR. The healthcare system is increasingly technology-dependent, and informatics skills are essential for HCPs to operate efficiently in the contemporary clinical environment (Raghunathan, McKenna, & Peddle, 2023). HCPs are pivotal for advancing digital services in healthcare settings (Nair, Kue, Athilingam, Rodríguez, & Menon, 2023). This study showed that most study participants had a positive perception of using mobile technology in health care. Indeed, the results generally prove that ECR allows for appropriate diagnosis and proper management of patients. It is necessary to combine the introduction of technology with support and management mechanisms (Blanchet, Sanon, Sarrassat, & Satouro Somé, 2023). This finding aligns with and reinforces the findings of Divall et al. (2013). For the latter, “Overall, healthcare professionals described using portable electronic devices, or personal digital assistants (PDAs) as a positive experience.” The added value of using PDAs is that they offer easy and quick access to health workers compared to paper-based devices, which are heavy and not easy to use. It is also important to highlight that managers’ attitudes play a great role in the success of the intervention: open dialogue and respect are crucial dimensions (Blanchet et al., 2023). Thus, the fact that health workers go through pages of paper protocols to obtain information, use several tables and guidelines and search for correct treatment doses made compliance with care protocols more complex. The ECR, therefore, appears in this context as a guide to diagnosis and treatment because it shows healthcare providers what questions to ask and what procedures to follow and often reminds them of what to do next. There is a need to ensure that healthcare organizations can use digital tools for service delivery (Morris et al., 2023).

In addition, the color of the classification is a central cue for HCPs using the ECR. The color of the classification, in addition to the classification itself, indicates to the HCPs the urgency and severity of the patient’s problem. This allows them to make management decisions quickly. The ECR combines the form and management guidelines developed by the MHPH. It will enable HCPs to have a single management tool without getting lost when filling out the form or using the management protocol. This is in line with the results of the study by Mitchell et al. (2013), particularly on using the ECR tool in the IMCI. This study revealed greater adherence to IMCI protocols when HCPs use electronic technology for care. Similarly, Bessat et al. (2019) showed in their study that the electronic device simplified the daily work of health staff, as all the assessment and management tools for children in one device are easier to use than the paper version.

Moreover, all the participants were willing to use the ECR. For us, they had a level of education (46% of participants had a general certificate of education at the ordinary level, and 48% had a general certificate of education at an advanced level or higher), which allowed them to understand and master the handling of the table.

Second, the training received by HCPs at the beginning of the implementation was an excellent strategy for increasing their buy-in. This is supported by Sawadogo et al. (2021), who mention that the expected organizational challenges include providing quality training to project participants, rigorous monitoring of mobile health activities by the implementation team, and well-defined tasks and measures to maintain participants’ motivation. Moreover, Cherrez-Ojeda et al. (2020) believe that adequate training of doctors in new technologies can significantly optimize digital resource use and work performance compared to those of untrained individuals. These skill enhancements allow health workers to acquire new competencies and break away from old practices considered more complex. Furthermore, the organization of care taught and the practical courses organized during these training sessions have enabled workers to redefine patient circuits and improve the care process. This makes it possible to reduce the waiting time for patients. Bessat et al. (2019) also confirmed that health workers perceive the ECR as a learning and training tool for the IMCI in that the device would increase their knowledge by helping them learn. Staff training (face-to-face or coaching) on digital tools was also crucial for successfully implementing and accepting the digital health strategy in the health districts.

Digital healthcare tools are becoming increasingly common worldwide, and similar issues have been identified across healthcare systems (Bondaronek, Dicken, Singh Jennings, Mallion, & Stefanidou, 2022). According to Mastellos et al. (2018), the degree of ICT used in health care considers the actors’ commitment but, more importantly, the hardware used. For example, when using ECR, the most common obstacles were application bugs. According to the authors, these application bugs led to the tablet’s slowness and recurrent breakdowns. In addition, there needs to be more staffing as a barrier when the size of the health staff is less than the volume of work to be done within a required timeframe. There is undoubtedly an increase in workload and a problem with synchronization.

Technical bugs malfunction when a health worker consults a patient. For example, an agent told us that when you reach a certain level during a consultation, the tablet takes you back to the beginning, and you are asked for the code to start the consultation again at zero. This situation led to discouragement on the part of the agents when faced with a patient who was waiting impatiently to be taken care of. This resulted in a longer waiting time for patients than usual. In addition, according to patients, these technical bugs created doubts and a feeling of incompetence on the agent’s part due to their fumbling with the tool. These concerns were reported by Perez et al. (2022) and Pędziński et al. (2013), who listed several barriers to implementing ICTs in healthcare, including technical barriers linked to software malfunction. Similarly, Bessat et al. (2019) reported that one of the barriers perceived by health workers was the slowness of the device, which was a source of complaint for some patients because it created longer waiting times.

Then, there are slow and recurrent crashes of the tablet. These differences were generally due to the tablets themselves. The slowness of the tablet is observed when switching from one form to another. Depending on the application used, the duration can vary from ten (10) seconds to two (2) minutes. This slowness could be linked to the low storage capacity and tablet quality. This leads to disappointment on the part of the agent who uses it and may lead him/her to file the ECR and move on with the paper version of the intake form.

The recurrent breakdowns of tablets include screen and charging problems. This can also lead to nonuse of the ECR and negatively influence the quality of the intake assessment. This leads us to agree with Haider et al. (2021) that in the context of using ICTs in the provision of care, the responsiveness of technical staff to user concerns is imperative for success. This provision could reduce the time needed to resolve queries, inventory the most recurrent problems, and set up a helpdesk to empower agents in dealing with similar issues. The problem of synchronization has been reported by Cherrez-Ojeda et al. (2020), who state that limited access to the Internet in mobile devices and work facilities is also considered a significant obstacle. This finding corroborates the synchronization problem identified in this study. An audit of the ECR tool could help *Foundation* Tdh technicians better understand the full scope of the application. In addition, the workload mentioned by some of the participants in our study and reported by Cherrez-Ojeda et al. (2020) could be linked to an organizational problem, as health workers cannot integrate tablets effectively into their routine organization.

Additionally, the Burkinabe health system still needs to review staffing standards at the HSPC level, integrating the use of digital tools in the activities of agents. This leads the latter to believe that the tablet has increased their workload even though they were using paper versions of these digital forms. This assertion is supported by Bessat et al. (2019), who mentioned in their study that several HCPs cited obstacles to adoption related to increased workload.

## Strengths and limitations

This study on the barriers and facilitators associated with using ECR for IMCIs is important because it is the first BF study. In addition, it focused on the very first health district to implement this technology in 2010 in BF. One of the strengths of this research lies in the careful choice of a sample consisting solely of key informants, in this case, HCPs, to gather the most significant possible wealth of information. This research underlines the relevance and enrichment of consulting HCPs directly involved in this technological implementation to have the best chance of shedding real light on the subject. The merit of this option is that it gave them a voice in highlighting the barriers and facilitating factors associated with using the ECR for IMCI, which was unknown in BF. The qualitative approach used in this study enabled us to highlight the barriers and facilitating factors associated with using ECR for IMCIs, which is not well known in BF. Although studies on ECR for IMCIs exist, the originality of this study is that it focused on HCPs as exemplary contributors to digital health for the benefit of child health. In addition, based on the experiences of HCPs analyzed in the context of a health district that has been in operation for more than a decade, this study provided a clearer picture of the issue. Therefore, lessons can be learned. Such a study is vital for improving the transferability of these materials. According to Yin (2009), this kind of study builds in-depth knowledge of a particular phenomenon so that the results can only be transferred to another group in the spirit of theoretical generalization. However, the results of this study, like all qualitative research, are only transferable to similar contexts where the ECR for IMCIs is used.

Additionally, one of the strengths of this study remains the contribution of information from three different data collection methods, namely, interviews and documentary reviews, which allowed for optimal triangulation. Other data sources corroborated all the statements collected from the HCPs. This triangulation ensures the validity and reliability of the data (Apostolidis, 2006).

Given the study’s limitations, there are undoubtedly limitations and potential biases in this study, even though precautions have been taken. We have limited the scope of our research to the sole point of view of HCPs. Although their point of view is inescapable and of great interest, we also felt it necessary and valuable to interview people who work closely with HCPs in implementing ECR in the HSPC. This would have provided a rich and relevant perspective on the research theme. Despite these limitations, this study highlighted the barriers and facilitators associated with using the ECR for IMCIs in the health district of Toma, BF.

## Conclusion

Using ECR for IMCIs is crucial for maintaining good-quality healthcare, especially for children under five in rural areas. The ECR for IMCIs can have long-term impacts on HCPs, children under five years of age, and the health system by improving data management for decision-making, the standard of healthcare service delivery and increasing attendance at health facilities and using services. Therefore, effective monitoring and evaluation of management changes before, during, and after ECR is used for IMCI are essential. Considering the potential barriers and facilitators of using ECR for IMCIs, all stakeholders should be informed, from planning until scaling up. In addition, it would be appropriate at the health facility level to consolidate facilitators and efficiently manage barriers with the participation of HCPs. Strengthening PHC facilities with qualified personnel and ensuring the stability of ECR applications are important for the easy use of ECR in IMCIs. Training and retraining HCPs are highly important when using ECR for IMCI. For a successful digital health strategy, it is advisable to focus on technical support and the quality of the technology. Above all, state-of-the-art electronic tools (updated versions) and a high-quality connection to the Internet network are essential.

## Data Availability

All data produced in the present work are contained in the manuscript

## Ethical considerations

The study received approval from the Burkina Faso Health Research Ethics Committee under deliberation N°2021-12-283. Investigation authorization was also obtained from the chief physician of the health district of Toma. Furthermore, the patients/participants provided written informed consent to participate in this study.

## Notes

### Competing Interest Statement

The authors have declared no competing interest.

### Funding Statement

This study did not receive any funding

### Author Declarations

The study received approval from the Burkina Faso Health Research Ethics Committee under deliberation number 2021-12-283. Investigation authorization was also obtained from the chief physician of the health district of Toma. Furthermore, the patients/participants provided written informed consent to participate in this study.

